# Addition of formaldehyde releaser imidazolidinyl urea and MOPS buffer to urine samples enables delayed processing for flow cytometric analysis of urinary cells

**DOI:** 10.1101/2022.04.07.22273579

**Authors:** Paul Freund, Christopher M. Skopnik, Diana Metzke, Nina Goerlich, Jan Klocke, Emil Grothgar, Luka Prskalo, Falk Hiepe, Philipp Enghard

## Abstract

Kidney diseases are a major health concern worldwide. Currently there is a large unmet need for novel biomarkers to non-invasively diagnose and monitor kidney diseases. Urinary cells are promising biomarkers and their analysis by flow cytometry has demonstrated its utility in diverse clinical settings. However, up to date this methodology depends on fresh samples, as cellular event counts and the signal-to-noise-ratio deter over time.

Here we developed an easy-to-use two-step preservation method for conservation of urine samples for subsequent flow cytometry. The protocol utilizes a combination of the formaldehyde releasing agent imidazolidinyl urea (IU) and MOPS buffer, leading to gentle fixation of urinary cells. The preservation method increases acceptable storing time of urine samples from several hours to up to 6 days. Cellular event counts and staining properties of cells remain comparable to fresh untreated samples.

The hereby presented preservation method facilitates future investigations on flow cytometry of urinary cells as potential biomarkers and may enable broad implementation in clinical practice.

## Introduction

Kidney diseases are among the most common diseases worldwide, affecting an estimated 10 % of the global population (1). Routine laboratory work-up of kidney diseases include serum creatinine, proteinuria and urinary sediment, which yield valuable information about kidney function and potential etiologies of functional impairment. However, the current parameters in use have only limited utility for establishing a diagnosis, predicting outcome and monitoring of treatment response. Kidney biopsy is the current diagnostic gold standard, but is invasive by nature and not free of risk, limiting its applicability especially in the follow up. Consequently, there is a large need for novel biomarkers, for non-invasive diagnosis, outcome prediction and monitoring treatment response.

Several groups including ours have reported flow cytometric analyses of urinary cell populations as non-invasive biomarkers in various settings, as it is already widely used for analysis of peripheral blood cells. Accordingly, increased numbers of urinary immune cell subsets, podocytes or tubular epithelial cells have been reported as biomarkers in several kidney diseases such as Lupus Nephritis (LN) (2-9), ANCA associated Vasculitis (10), Kidney Allograft Rejection (11), idiopathic Membranous Nephropathy (12), autosomal dominant polycystic kidney disease (13) and Acute Kidney Injury (14).

However, up to date flow cytometry of urine cells strictly depends on fresh samples, as prolonged incubation of cells in urine induces cell death and alters cell characteristics (15). This dependence on fresh samples makes this diagnostic approach expensive and limits its use to large hospitals with the required laboratory infrastructure. On-site conservation of cells could overcome these limitations and allow broader applicability. Here we report a user-friendly protocol utilizing a combination of pH-buffer and formaldehyde-releasing agent, enabling conservation of cells directly in the urine sample. The simple and robust two-step method can easily be performed in less than a minute and enables broader clinical application.

## Material and Methods

### Patient cohort

To investigate the influence of the chemical agents on cells, especially regarding staining quality, we used urine spiked with PBMCs from healthy donors. PBMCs were isolated as described before (16). To validate our preservation method as shown in Figures 3 and 4, we collected urine samples of 17 Patients in November and December 2020 from the Department of Nephrology, Charité University Hospital, Berlin. We included patients with high probability for a significant number of urinary cells regarding to their clinically suspected or diagnosed kidney disease: Deterioration of Kidney Transplant Function (n = 8), Urinary Tract Infection/Pyelonephritis (n = 6), Acute Interstitial Nephritis (n = 1), Glomerulonephritis (n = 2).

All patients and healthy donors gave their informed consent for study participation and scientific publication of the results. This study was approved by the ethics board of Charité University Hospital, Berlin (approval number EA1/284/19).

### Sample collection and preparation

Within 3 hours after voiding urine samples were collected and processed. Samples were split into equal parts to allow direct comparison of different storage conditions. Storage conditions and respective incubation times are indicated in the results section for each experiment.

Minimum urine volume was 40 ml, mean urine volume was 68 ml. From all 17 urine samples four samples were collected from urinary catheters. For fixation of samples, we added liquid MOPS (1 mol/L) in a 1:3 proportion of total volume and powdery imidazolidinyl urea (20 g/L of total volume) (Sigma-Aldrich) and stored them for assigned time at 4 °C, if not indicated otherwise, prior to staining and flow cytometric analysis.

### Composition and usage of MOPS buffer (1 mol/L)

20.95 g 3-(Morpholin-4-yl) propane-1-sulfonic acid (MOPS) (Carl Roth), 0.82 g sodium acetate (Carl Roth), 1.85 g Ethylenediaminetetraacetic acid (EDTA) (Sigma-Aldrich) were dissolved in 100 ml deionized water and adjusted to pH 7 with sodium hydroxide (Carl Roth). This was used as a 3x-solution in urine specimen.

### Staining and flow cytometry

After centrifugation (600 x g, 8 min, 4 °C) and resuspension of the resulting pellet in phosphate-buffered saline (PBS)/bovine serum albumin (BSA)/Ethylenediaminetetraacetic acid (EDTA) we divided each sample for staining into a leukocyte panel and an epithelial cell panel, each with a corresponding staining control. In the leukocyte panel we stained for T cells (CD3+) and the subpopulations T helper cells (CD3+CD4+) and cytotoxic T lymphocytes (CD3+CD8+), as well as monocytes/macrophages (CD14+CD36+). In the tubular epithelial cell panel, we detected epithelial cells using an anti-cytokeratin-antibody, followed by a staining for CD10+ and/or CD10+CD13+ epithelial cells (indicating proximal tubular epithelial cells (17, 18) and CD326+ epithelial cells (CD326 is also known as EpCAM, indicating distal tubular epithelial cells (19, 20).

We used the following monoclonal antibodies for staining: CD4-PE-Vio770 (clone: REA623), CD14-FITC (clone: TüK4, isotype mouse IgG2a), CD36-PE (clone: AC106, isotype mouse IgG2a), Cytokeratin-FITC (clone: CK3-6H5, isotype mouse IgG1), CD10-PE-Vio770 (clone: 97C5, isotype mouse IgG1), CD13-APC-Vio770 (clone: REA263), EpCAM-PE (clone: HEA-125, isotype mouse IgG1) (all Miltenyi Biotec), CD3-eFlour780 (clone: SK7, mouse IgG1) (eBioscience), CD8-Alexa647 (clone: GN11/134D7, mouse IgG1) (DRFZ).

In fresh samples we fixed and permeabilized cells with BD Cytofix/Cytoperm solution (BD Biosciences) for intracellular staining of cytokeratin (only in epithelial cell panel).

Prior to incubation with the antibody mix for 15 min at 4 °C in the dark we blocked unspecific binding of antibodies with 10% human IgG (Gammunex, Grifols) added in a 1:100 ratio to PBS/BSA/EDTA (Leukocyte Panel) or Perm/Wash-buffer (BD Biosciences) (Epithelial Cell Panel), respectively.

We performed flow cytometric analysis with BD FACSCanto II (BD Biosciences) according to previously published guidelines (21) after obtaining a single-cell suspension using a 30 µm pore size cell strainer.

### Cryoconservation of samples

For freezing experiments, we treated urine samples with the preservation method for one day prior to centrifugation (600 x g, 8 min, 4 °C) and resuspension of the resulting pellet in freeze medium (DRFZ, Berlin, fetal calf serum + 10 % DMSO). These samples were stored at −80 °C for 28 days. After thawing, we performed a rinsing step, i.e. centrifugation and resuspension of the resulting pellet in PBS, before proceeding to the staining process as described above.

### Statistical analysis and data plotting

We calculated Pearson’s correlation coefficients and corresponding significance levels using student’s t-tests with R (v4.1.0) / RStudio (v.1.4.1717). For analysis we used the following packages: flowCore (v.2.4.0), flowWorkspace (v.4.4.0) and CytoML (v.2.4.0) for importing flow cytometry data previously analyzed with FlowJo (v.10.7.1, FlowJo, BD Biosciences), tidyverse (v.1.3.1, including ggplot, tidyr, dplyr, tibble, stringr), scales (v.1.1.1) and cowplot (v.1.1.1) for plotting data sets.

## Results

### Cellular event counts and staining quality decrease over time in untreated urine samples

We stored untreated patients’ urine samples at 4 °C and analyzed the sediments with flow cytometry at predefined time points, immediately after voiding (fresh) and after 1, 3 and 6 days. Over time, we observed an altered staining quality with reduced signal-to-noise ratios for several analyzed epitopes (*Fig 1A*) and a decrease of absolute cellular event counts *(Fig 1B)*.

**Fig 1.**
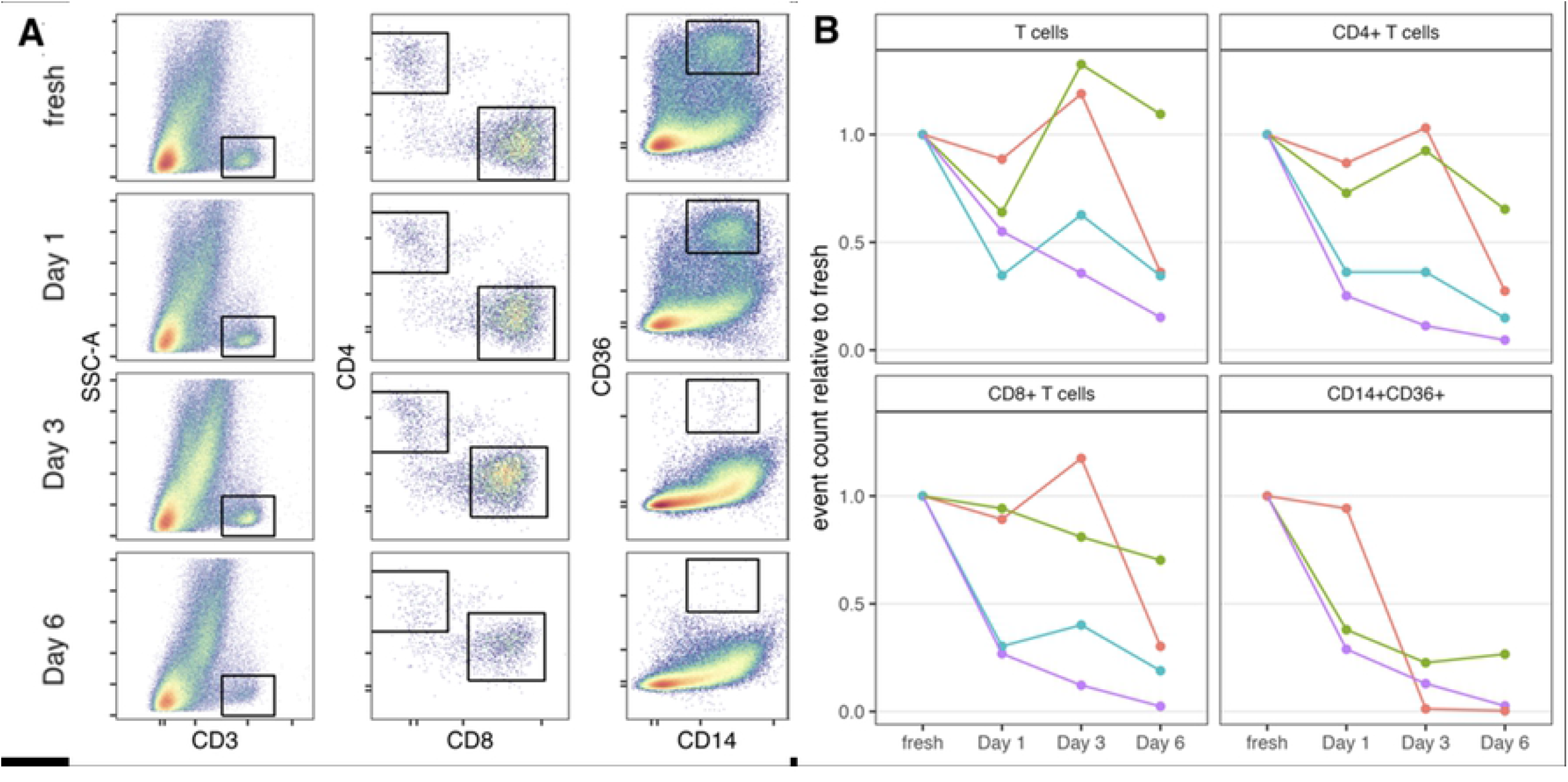
Storage of untreated urine samples at 4 °C reduces staining quality and cellular event count. Loss of detectable immune cells by flow cytometry is observed after incubation in urine over time. (A) Representative pseudocolor plots with gates for CD3+ T cells, CD3+CD4+ and CD3+CD8+ T cells and CD14+CD36+ monocytes/macrophages after various incubation times in urine. Prior gates for lymphocytes and doublet exclusion in scatter dimensions are not shown; Live/dead discrimination was not performed. (B) Respective relative event counts of gates in A for multiple urine samples (color coded). Cellular event counts are displayed as events in relation to events measured immediately after voiding (fresh).

### Addition of formaldehyde to urine samples elicits precipitates and reduces binding of antibodies to their epitopes

Fixation of cells and tissues with formaldehyde is routine practice in laboratories. Therefore, we next assessed whether this could be applied to urine samples. Addition of formaldehyde (1 %) directly to patients’ urine specimen caused precipitation of a gel-like mass *(Fig 2A)* in 14 of 26 samples after approximately 20 hours at room temperature. This prohibited analysis by flow cytometry. Dipstick testing revealed that precipitate formation was more likely in samples with low pH and increased specific gravity *(Fig 2B)*.

**Fig 2.**
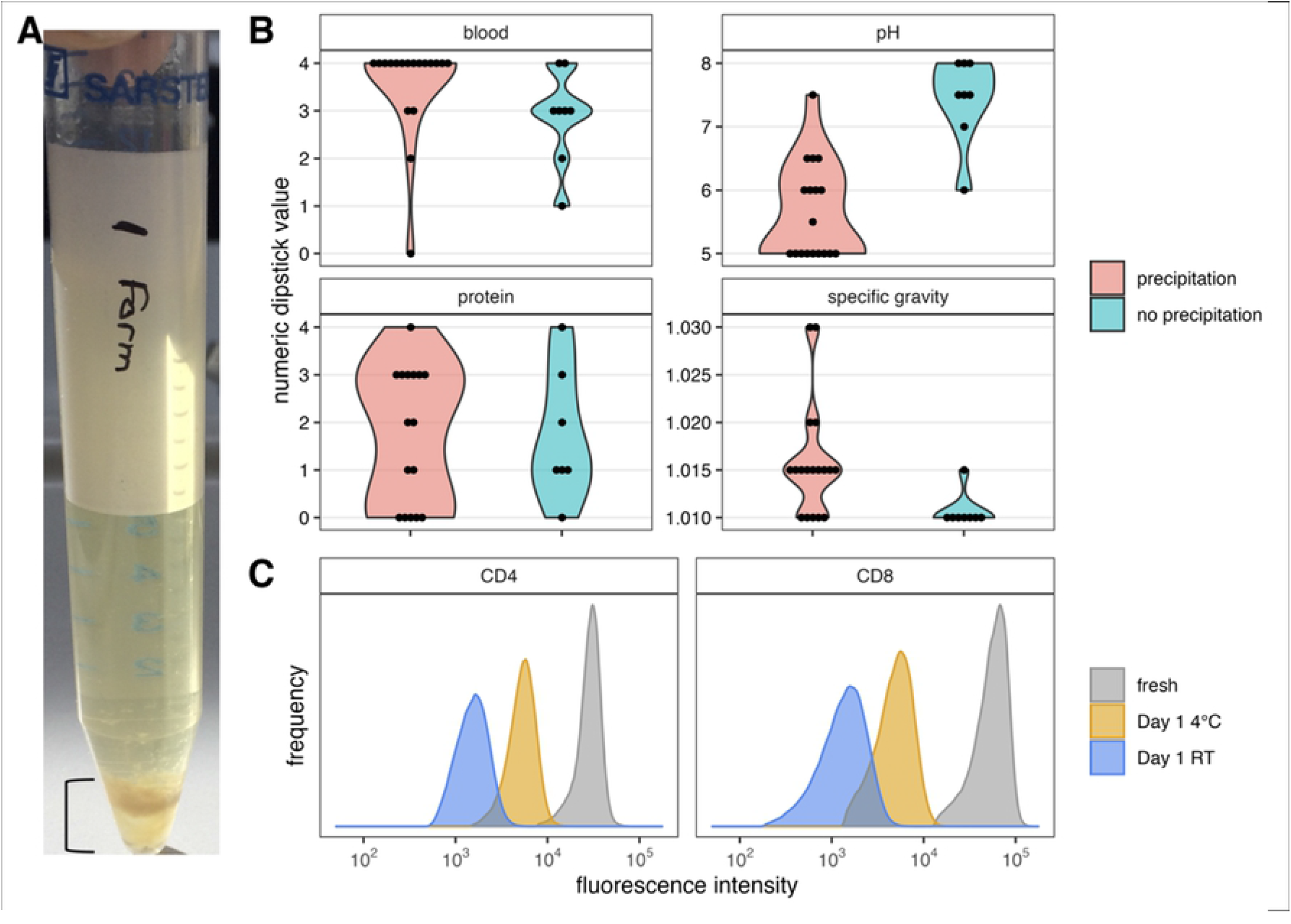
Formaldehyde as a fixative in stored urine samples causes precipitate formation and reduces antibody binding. (A) Example of precipitate in patients’ unbuffered urine incubated with 1 % formaldehyde formed after 1 day. (B) Comparison of urine sample characteristics and precipitate formation in patients’ urine incubated with 1 % formaldehyde for 20 hours at room temperature (RT). Note the effect of low pH and high specific gravity. (C) Fluorescence intensities of T cell surface antigens labeled with fluorochrome-conjugated monoclonal antibodies. Fluorescence intensities decrease after 1 day of incubation of PBMCs in urine treated with MOPS and formaldehyde. Storage at room temperature (RT) increases the effect.

In order to buffer urine specimen close to pH 7 we added dissolved, highly concentrated 3-(Morpholin-4-yl) propane-1-sulfonic acid (MOPS, buffer range pH 6.5 - 7.9) buffer adjusted to pH 7 with sodium hydroxide (see Material and methods). This prevented precipitation in subsequent samples. Next, we incubated human peripheral blood mononuclear cells (PBMCs) in urine and added formaldehyde and MOPS buffer for at least 20 hours. Subsequent flow cytometry analysis revealed gross reduction of mean fluorescence intensities after staining with fluorochrome-conjugated monoclonal antibodies, indicating significant loss/alteration of surface epitopes *(Fig 2C)*.

### Treatment of urine samples with imidazolidinyl urea (IU) and MOPS buffer allows detection of urinary cells for up to six days

As an alternative fixation strategy aiming at a gentler fixation we next tested the formaldehyde releaser imidazolidinyl urea (IU). A cohort of patients’ urine specimen expected to contain high numbers of cells were treated with IU and MOPS buffer and stored for up to six days at 4 °C. Urine sediments were then analyzed by flow cytometry using two panels of well-established monoclonal antibodies – one for leukocytes and one for kidney tubular epithelial cells (TEC). No loss of staining intensity was observed despite storage for 6 days *(Fig 3)*.

**Fig 3.**
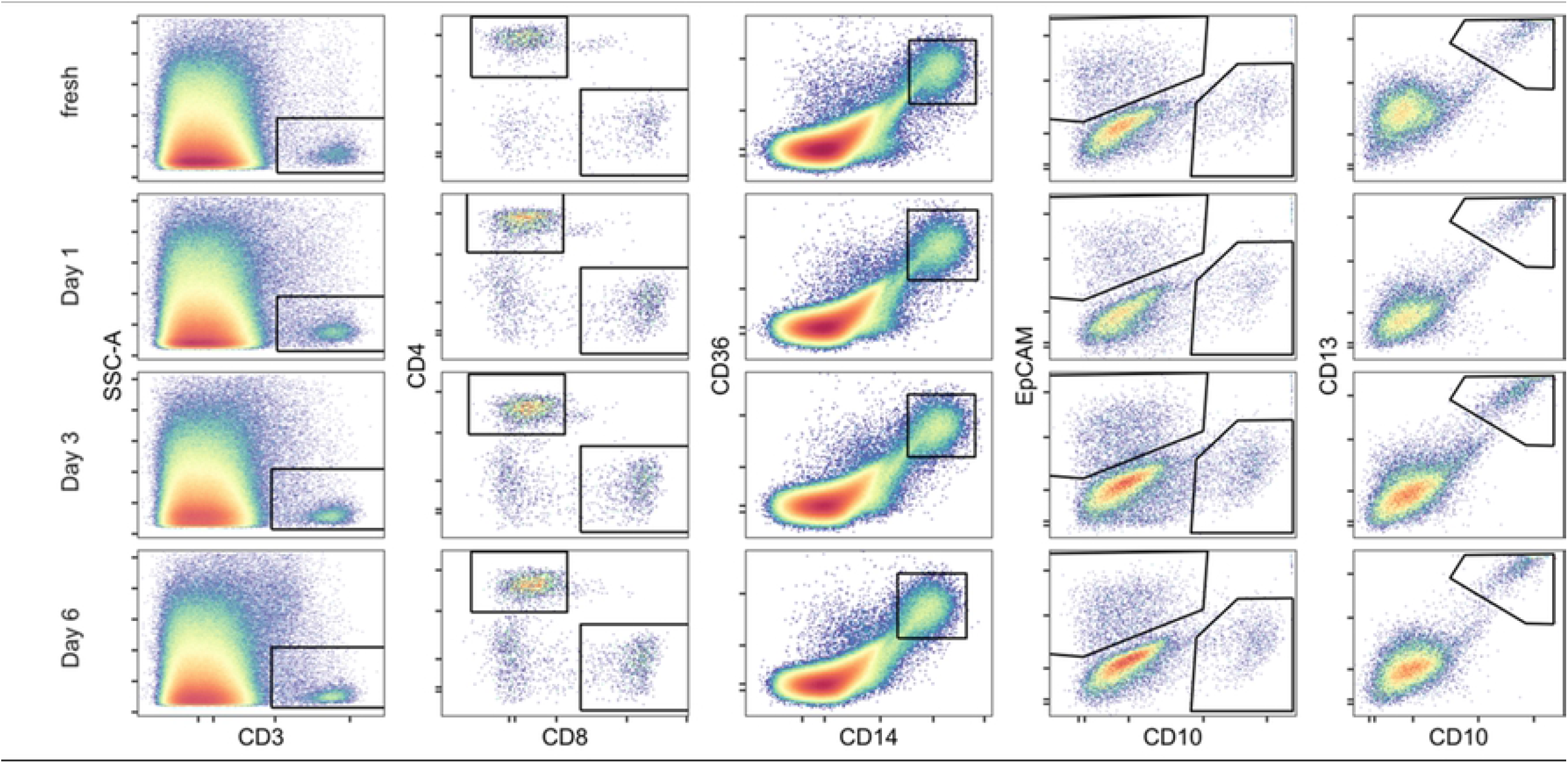
Treatment of urine samples with IU and MOPS buffer preserves urinary cellular events for up to one week. Analyses of leukocytes (column 1-3) and tubular epithelial cells (column 3-4) derived from preserved urine samples after different timepoints (rows 2-4) are highly comparable to fresh samples (row 1). Prior gates for exclusion for doublets and leukocyte gate or exclusion of debris and gating for cytokeratin+ epithelial cells (tubular epithelial cells), respectively, not shown.

### Cellular event counts remain stable despite fixation and storage for up to six days

Next, we investigated whether fixation and storage of urine samples would alter the respective cellular event counts in the samples. To this end, the quantity of cellular events in fresh samples was compared to fixed samples on different time points. Cellular event counts derived from preserved urine samples showed excellent correlation with cellular event counts derived from fresh samples (R ≥ 0.85) for all analyzed cell types and on all assessed time points *(Fig 4)*. Even after storing preserved urine samples for 6 days, we could demonstrate a strong consistency in detected leukocyte counts: T cells (CD3+, R = 0.88 (CI 0.70, 0.96), p < 0.001, *not shown*), T helper cells (CD3+CD4+, R = 0.94 (CI 0.84, 0.98), p < 0.001, *Fig 4, column 1*), cytotoxic T lymphocytes (CD3+CD8+, R = 0.94 (CI 0.83, 0.98), p < 0.001, *not shown*) and monocytes/macrophages (CD14+CD36+, R = 0.93 (CI 0.82, 0.98), p < 0.001, Fig 4, *column 2*).

**Fig 4.**
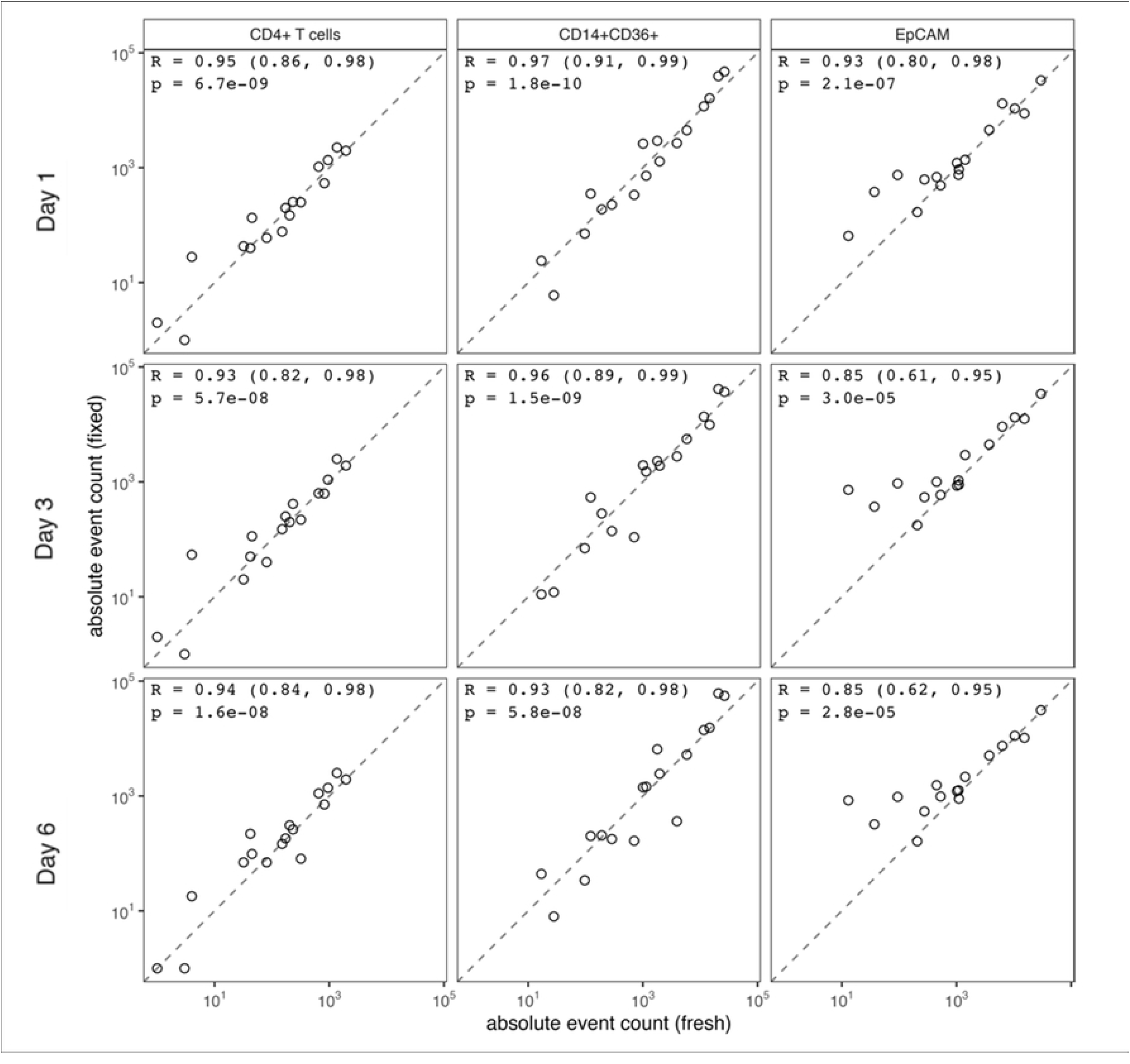
Correlation of cellular event counts derived from fresh samples with fixed samples after one, three and six days. Cellular event counts of CD4+ T helper cells, monocytes/macrophages (CD14+CD36+) and distal tubular epithelial cells (Cytokeratin+EpCAM+) are displayed. Dashed lines indicate ideal correlation. Pearson’s correlation coefficient (R), and p-value displayed.

Although correlation coefficients of tubular epithelial cells were comparable to leukocytes after 1 day, correlation coefficients decreased slightly on later time points, whilst still indicating a strong correlation after 6 days: distal tubular epithelial cells (EpCAM+, R = 0.85, (CI 0.62, 0.95), p < 0.001, *Fig 4, column 3*), CD10+ proximal tubular epithelial cells (CD10+, R = 0.85 (CI 0.61, 0.95), p < 0.001, *not shown*), CD10+CD13+ proximal tubular epithelial cells (CD10+CD13+, R = 0.87 (CI 0.66, 0.95), p < 0.001, *not shown*).

### Cryoconservation of IU/MOPS treated urinary cells

Aiming to increase intervals between urine voiding to processing and analysis even further, we stored patient samples treated with the described preservation method at −80 °C for 28 days. After freezing and storage at −80 °C, the flow cytometric staining quality and the quantity of cellular events remained comparable to samples analyzed one day after specimen collection *(Fig 5)*. This was also observed for cell populations not displayed: T cells (CD3+, R = 0.96 (CI 0.50, 1.00), p < 0.05), cytotoxic T lymphocytes (CD3+CD8+, R = 0.97 (CI 0.63, 1.00), p < 0.01), CD10+ proximal tubular epithelial cells (CD10+, R = 0.93 (CI 0.26, 1.00), p < 0.05), CD10+CD13+ proximal tubular epithelial cells (CD10+CD13+, R = 0.93 (CI 0.27, 1.00), p < 0.05).

**Fig 5.**
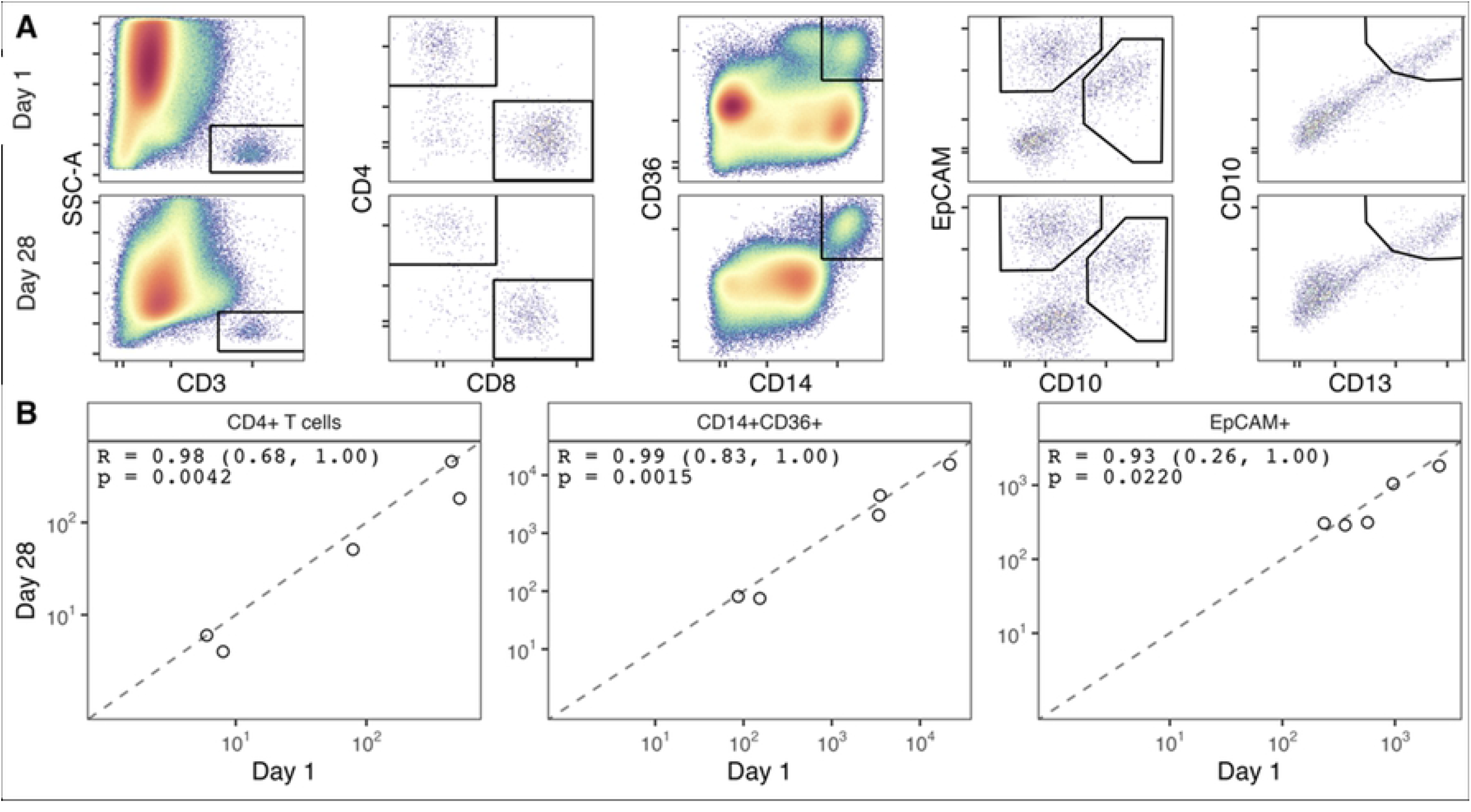
Staining quality and cellular event counts remain stable when fixed samples are cryoconserved for 28 days. (A) Comparison of pseudocolor plots of different analyzed cell populations; freezing and storage for 28 days at −80 °C does not change staining properties (top row: fixed cells, immediately analyzed after 1 day; bottom row: fixed cells, analyzed after 28 days at −80 °C). (B) Storage at −80 °C for 28 days does not change quantity of cellular events. (Dashed line indicates ideal correlation; Pearson’s correlation coefficient (R) and p-value displayed)

## Discussion

Analysis of urinary cells using state-of-the-art technology such as flow cytometry holds great potential for substantially improving the diagnosis and monitoring of kidney diseases. In order to facilitate such an analysis we here report a simple protocol for conservation of urinary cells.

Most studies investigating flow cytometry of urinary cells as biomarkers relate on cellular event counts in some way. Therefore, two key requirements need to be met when establishing a method for conserving urinary cells for subsequent flow cytometry: preservation of epitopes for detection with antibodies and consistency in cellular event counts.

Formaldehyde is widely known in the medical field as a fixative for cells and tissues, as it prevents degradation of cells by cross-linking proteins on cell surfaces (22). Adding formaldehyde as a fixative agent directly to urine samples however failed to meet the above stated requirements. First, formation of precipitates upon incubation was observed, prohibiting further analysis. We assume that these precipitates are glycoprotein uromodulin (also known as Tamm-Horsfall-Protein), a protein excreted into urine in considerably quantities by tubular epithelial cells of the kidney. This assumption is supported by the observation that especially urine samples with low pH tended to form precipitation, as precipitation of uromodulin is favored in low pH environments and high osmotic concentrations (23). Furthermore, fixation of urinary cells with formaldehyde resulted in alteration/loss of surface epitopes. This impeded identification of cell populations upon staining with respective antibodies, which is a prerequisite for flow cytometry. We believe the loss/alteration of epitopes to be the result of overfixation, as storage at room temperature further deteriorated the staining quality.

As alternative to formaldehyde we subsequently used imidazolidinyl urea as a fixative - a powdery formaldehyde-releasing agent commonly used in cosmetics as a preservative (24). It slowly releases formaldehyde upon dissolving, which then acts as described above. Using imidazolidinyl urea led to fixation of cells yet preserving epitopes for specific detection.

MOPS (3-(N-morpholino)propanesulfonic acid) is a buffer first introduced by Good et al in 1972 for biochemical studies (25). MOPS offers the advantages of good water solubility, inertness in biological/biochemical reactions and low/no cell membrane permeability, making it an ideal buffer candidate in our setting. Addition of MOPS to the urine samples prevented formation of precipitates.

As presented here, our method increases the acceptable interval between sample collection and definitive processing from several hours to up to 6 days. Our protocol achieves both requirements mentioned above with relatively cheap chemicals and without any expensive equipment. By freezing preserved samples, this interval can be extended even further. Therefore, more samples can be analyzed as a batch, facilitating standardization in processing and ultimately increasing the reliability of results.

Importantly, our method consists of a simple two-step protocol - a predefined volume of urine is added into a prepared sample cup containing buffer prior to adding the formaldehyde releaser (imidazolidinyl urea, IU). Thus, it can be applied easily even in a time sensitive setting such as the clinical context and it does not require any special laboratory training.

Developing a one-step method for the addition of chemical agents could even further improve and simplify preanalytical handling of urine in the future. Additionally, testing the combination of our preservation method with sample freezing at −80 °C for even longer time intervals could benefit long term storage possibilities. Lastly, more urinary cell types should be examined for compatibility with our protocol; especially podocytes, which have been reported as biomarkers previously (12) (11).

In summary, our conservation method makes flow cytometry of urinary cells more accessible to future research, as it facilitates sample logistics and prolongs storage intervals. Without interfering with staining quality and quantity of detectable cell counts our protocol enables user-friendly sample processing and storage for up to six days, and even longer if samples are frozen. We believe this brings us one step closer to establishing urine cell analyses as biomarkers in clinical practice.

## Data Availability

All fcs files will be made available from FlowRepository after acceptance. R code will be made available from GitHub after acceptance.

## Supplemental

**S1 Gating strategy of urinary cell flow cytometry** Doublet exclusion in Forward and Sideward Scatter Channels (FSC, SSC, respectively) was followed by a broad leukocyte scatter gate. Monocytes were identified via CD14 and CD36 double positivity and subsequent back-gating to FSC and SSC. T cells were identified via CD3 staining, followed by a scatter back-gating to the FSC and SSC gate and finally by discrimination between CD4+ and CD8+ T cells.

**S2 Gating tree displaying the used gating strategy**

